# Long term effectiveness of inactivated vaccine BBIBP-CorV (Vero Cells) against COVID-19 associated severe and critical hospitalization in Morocco

**DOI:** 10.1101/2022.01.25.22269822

**Authors:** Jihane Belayachi, Majdouline Obtel, Rachid Razine, Redouane Abouqal

## Abstract

**INTRODUCTION:** We provide national estimates of the real-world Vaccine effectiveness (VE) based on nationally available surveillance data. The study aimed to estimate the effectiveness of the inactivated Covid-19 vaccine BBIBP-CorV (Vero Cells) Sinopharm vaccine currently deployed in Morocco to reduce the risk of hospitalization from a severe infection of SARS- CoV-2 virus within 9 months after vaccination.

**METHODS:** We conducted a test-negative, case-control study among a population aged 18 years or older who were tested by rt-PCR for SARS-CoV-2 infection from February to October 2021 in Morocco. From the national laboratory COVID-19 database; we identified cases who were rt-PCR positive amongst severe and critical COVID-19 cases and controls who had a negative rt-PCR test for SARS-CoV-2. From the national vaccination register (NVR); individuals vaccinated with COVID-19 Vaccine (Vero Cell) and those unvaccinated were identified and included in the study. The linkage between databases was conducted for the study of Vaccination status based on the timing of the vaccine receipt relative to the SARS-CoV-2 rt-PCR test date. For each person, who tested positive for SARS-CoV-2, we identified a propensity score-matched control participant who was tested negative. We estimated vaccine effectiveness using conditional logistic regression.

**RESULTS:** Among 12884 persons who tested positive and 12885 propensity score-matched control participants, the median age was 62 years, 47.2% of whom were female. As a function of time after vaccination of second dose vaccination, vaccine effectiveness during the first month was 88% (95% CI, 84-91), 87% (95% CI: 83-90) during the second and third month, 75% (95% CI: 67-80) during the fourth month, 61% (95% CI: 54-67) during the fifth month, and 64% (95% CI: 59-69) beyond the sixth month. VE remained high and stable during the first three months in the two-age subgroup. In the fourth month, the VE in the older population aged 60 years and above (64%) was reduced by 20 points compared to VE in the younger population (84%).

**CONCLUSION:** A Sinopharm vaccine is highly protective against serious SARS-CoV-2 infection under real-world conditions. Protection remained high and stable during the first three months following the second dose and decreases slightly beyond the fourth month especially beyond 60 years.

## INTRODUCTION

As of January 24, 2022, the COVID-19 pandemic has resulted in more than 330 million cases and more than 5.5 million deaths worldwide, including 1.06 million cases and 151⍰025 deaths in the Kingdom of Morocco ^1^.

The development and deployment of sustainably effective vaccines amongst susceptible and at-risk populations remain key to end the pandemic. Since January 2021, four COVID-19 vaccines - BNT162b2 (Pfizer-BioNTech), Ad26.COV2.S (Johnson & Johnson-Janssen), the ChAdOx1 nCoV-19 (AZD1222; Oxford-AstraZeneca) and BBIBP-CorV (Vero Cells) Sinopharm; received emergency use authorization in Morocco based on short-term safety and efficacy against COVID-19 and were deployed in Morocco shortly thereafter.

In January 2021, a nationwide vaccination campaign was launched in Morocco; to administer COVID-19 vaccines for people aged 18 years and above. As of January 2022, one year after the start of the vaccination campaign it is estimated that nearly 70% of the target population (30,000,000) has received at least one dose of vaccine, with Sinopharm amongst 72% of vaccine receipt ^2^.

Phase III trials reported high vaccine efficacy against SARS-CoV-2 infection with, two doses of the inactivated COVID-19 vaccine showing 72.8% and 78% efficacy against symptomatic coronavirus disease 2019 and 79% (26-94) efficacy against severe disease or hospitalisation 21 days after the first dose in people aged 18 years and above. However, Phase III trials data provided sparse information on the impact of vaccination on transmission of SARS-CoV-2 infection and did not take into account different population groups; and data shown are on protection against SARS-CoV-2 ancestral strain.

Since the start of the outbreak, Morocco has experienced three epidemic waves, the second wave (from July to September 2021) noted the dominance of circulation of Delta (B.1.617.2) variant (80%). The current third wave is characterized, during early January 2022, by a circulation of Omicron (B.1.1.529) variant predominantly (70%). Given the short time since the introduction of the COVID-19 vaccine, the short duration of follow-up may have underestimated the efficacy of the vaccine and their duration of protection against serious consequences, which will be important to inform treatment strategies, administration of booster doses, and assessing the evolution of vaccine efficacy with the emergence of new VOCs. Yet, data are limited on the real-world effectiveness of inactivated COVID-19 vaccine BBIBP-CorV (Vero cells). The effectiveness and duration of protection offered by the inactivated COVID-19 vaccine BBIBP-CorV (Vero cells) against serious or critical hospitalizations associated with COVID-19 are not known in Morocco.

We provide national estimates of the real-world effectiveness based on surveillance data available in the Kingdom of Morocco, which has a population of approximately 37 million inhabitants. This study aimed to estimate the effectiveness of the inactivated COVID-19 vaccine BBIBP-CorV (Vero Cells) Sinopharm currently deployed in the Kingdom of Morocco to reduce the risks of severe hospitalization associated with SARS-CoV-2 infection 9 months after completing the vaccination.

## Methods

### Study design

We estimate the real-world vaccine effectiveness (VE) over time after administration of two doses of inactivated COVID-19 vaccine BBIBP-CorV (Vero Cells) Sinopharm against COVID-19-associated severe or critical hospitalized cases using a test-negative, case- control study design comparing the odds of a positive SARS-CoV-2 test result between vaccinated and unvaccinated patients.

### Study population

Eligible medical encounters were defined as those among adults aged 18 years and above who had a reverse transcription real-time polymerase chain reaction (rt-PCR) test for SARS-CoV-2 infection between February 01 2021 and October 01 2021. The start of the study period corresponded to 14 days after the first individuals received the first dose of Sinopharm vaccine (index date). The study period started on February 12, 2021, 14 days after receiving the first dose of the Sinopharm COVID-19 vaccine. Recipients of BNT162b2 (Pfizer-BioNTech), Ad26.COV2.S (Johnson & Johnson-Janssen), the ChAdOx1 nCoV-19 (AZD1222; Oxford-AstraZeneca, and those for whom 1–13 days had elapsed since receipt of the first dose Sinopharm COVID-19 vaccine were excluded.

### Exposure

Vaccination status was categorized based on the number of vaccine doses received and a number of days from vaccination to the sars cov 2 rT PCR.

- Unvaccinated
- Partial vaccination: included patients who received only the first dose of the vaccine and were tested 14 days and more after the first vaccination.
- Any time after 2^nd^ dose: Included individuals with receipt of a second dose of vaccine regardless of days before the index date.

Collected data included age and gender, rt-PCR test calendar date, rt-PCR test results, geographic location, and the 7-day moving average of the percentage of SARS-CoV-2 rt-PCR positive tests. The 7-day moving average of the percentage of SARS-CoV-2 rt-PCR positive tests was extracted from public health records; as a measure of SARS-CoV-2 circulation on the day of each rt-PCR test. Infection and vaccination statuses were both ascertained at the time of the PCR test.

### Data sources

The following data sources were used:

Vaccination histories were obtained from a common, unbiased electronic health record system. This dataset was based on a comprehensive and inclusive population-based list of target populations drawn up firstly for the entire population over the age of 17 based on a national identity card (NIC). This list is the basis of the national vaccination register (NVR). Vaccine receipt is associated with the registration of vaccine information in the NVR. The date when the COVID-19 vaccine generally became available, according to the age group of the recipients, was provided. Vaccine information included (vaccine date, vaccine type and vaccine dose).

E-labs is a national laboratory COVID-19 network for diagnostic specimens tested by Reverse Transcription Polymerase chain reaction (rt-PCR) in Morocco. Individuals who were tested for SARS-CoV-2 by rt-PCR were potential subjects for this study. The test result by rt-PCR for all individuals tested and included in this study was obtained^3^. COVID-19 test result for individuals based on rapid antigen-based test kits or those clinically diagnosed is not registered in the E-labs registry. Clinical data of all COVID-19 associated severe or critical hospitalisation are registered in a minimal dataset made for public health surveillance.

National identity card number is used as the key to linking the three databases (NVR and E-labs registry and clinical minimal dataset).

### Study groups

The case group were subjects with severe or critical infection with SARS-CoV-2 infection confirmed by the rt-PCR positive test result. The control group were individuals who were tested negative for SARS-CoV-2 infection on an rt-PCR test.

- For each case, we considered the first rt-PCR -positive test for SARS-CoV-2 infection with associated COVID-19-related severe or critical hospitalisation during the study period and we excluded all those who were tested negative for SARS-CoV-2 infection. For each control, we considered the first rt-PCR-negative test for SARS-CoV-2 infection during this period. This strategy was used to control for potential bias due to repeat testing in rt-PCR-positive individuals seeking to check for infection clearance or bias arising from repeat testers among controls (persons with a higher level of healthcare-seeking behaviour and presumably lower risk of infection).
- Severe and critical COVID-19 was defined according to WHO definition^4^

### Ethical Considerations

This real-world study was pre-registered (https://osf.io/at3yf) on Open Science Framework. The study was approved by the Rabat local ethic committee review board for biomedical research at Mohammed V University (N/21). A waiver of informed consent was granted for the study. Authorization N° A-RS-638/2021 from the National Commission for the Protection of Personal Data (CNDP) was obtained.

### Sample size

Although studies with large national datasets do not need to calculate the minimum sample size^5^, we calculated the minimum sample size, considering that subgroup analysis will be needed to evaluate the vaccine protection effect (VE) for severe or critical infections. We estimated that VE is 80%, the population vaccination rate is approximate 50%. If the VE estimation accuracy is ±5%, and α=0.05, according to the 1:1 matching of cases and controls, 2768 cases and 2768 controls are needed. Thus, 2768 in each of the two groups, with a total of 5536 subjects.

### Statistical analysis

We conducted a univariate analysis of SARS-CoV-2–positive patients with negative controls. A standardized mean or proportion difference of more than 0.2 was considered to be noteworthy^6^.

We performed the iterative propensity score logistic regression model search procedure described by Imbens and Rubin. Given a binary outcome measure and a list of explanatory variables (sex, age, calendar days of the rt-PCR test; geographic location; and the 7-day moving average of the percentage of SARS-CoV-2–positive test), we iteratively selected explanatory variables for inclusion into a propensity score model based upon changes in the logistic regression log-likelihood. We also produced multiplicative interactions (and second-order terms) of all explanatory variables selected for model inclusion and treat interaction terms as additional candidate covariates^7^.

We matched cases and controls in a one-to-one ratio by propensity score^8^.

We used nearest-neighbour matching with a calliper of 0.2SD. We estimated vaccine effectiveness using conditional logistic regression to calculate the odds of testing positive among the vaccinated group versus the unvaccinated group^9^.

Vaccine effectiveness (VE) and its associated 95% CI were then estimated using the formula VE = (1 − Odds Ratio of vaccination among cases versus controls) x100%.

We evaluated the protective efficacy of vaccine on the people with serious, critical symptoms and positive for SARS-COV-2 by rt-PCR with:

1. Partial schedule vaccination (14 days or more after administration of the first dose).
2. Anytime schedule vaccinated (from 1st day to 9th months after the administration of second dose vaccines).
3. Overtime after second dose receipt, by range time including:

(a) From 1st day to 30th days (during the first month after the second dose); (b) From 31th to 60th days (during the second month after the second dose); (C) From 61th to 90th days (during the third month after the second dose); (d) From 91st to120th days; (e) From 121th to 150th; and (f) beyond 150th day to 9th month.

Additional analyses were performed to estimate vaccine effectiveness stratified by age (1⍰ 60 and > 60 years) and sex any time after the second dose, and in three range time to determine vaccine effectiveness in various population subgroups. Effectiveness was calculated for each subgroup. For all point estimates of vaccine effectiveness, we calculated 95% CIs ^10^.

All the analyses were conducted using Stata (version 14; StataCorp, College Station, TX, USA) and R software, (version 4.1.2; R Foundation for Statistical Computing).

## RESULTS

Among 12884 persons who tested positive and 12885 propensity score-matched control participants, the median age was 62 years. We identified 25,768 matched pairs of patients who were tested for SARS-CoV-2 infection between 1 February 2021 and 01 October 2021 (1:1 ratio of positive and negative test results for SARS-CoV-2 infection). The median age was 62 years among those who tested negative and 63 years among those who tested positive for SARS-CoV-2 infection; in both groups, 47.2% of participants were female. Among those who tested positive, 1015 (7.9%) had been vaccinated, and among those who tested negative, 2935 (22.8%) had been vaccinated. Baseline characteristics of the study population are shown in Table 1.

**Table 1:**
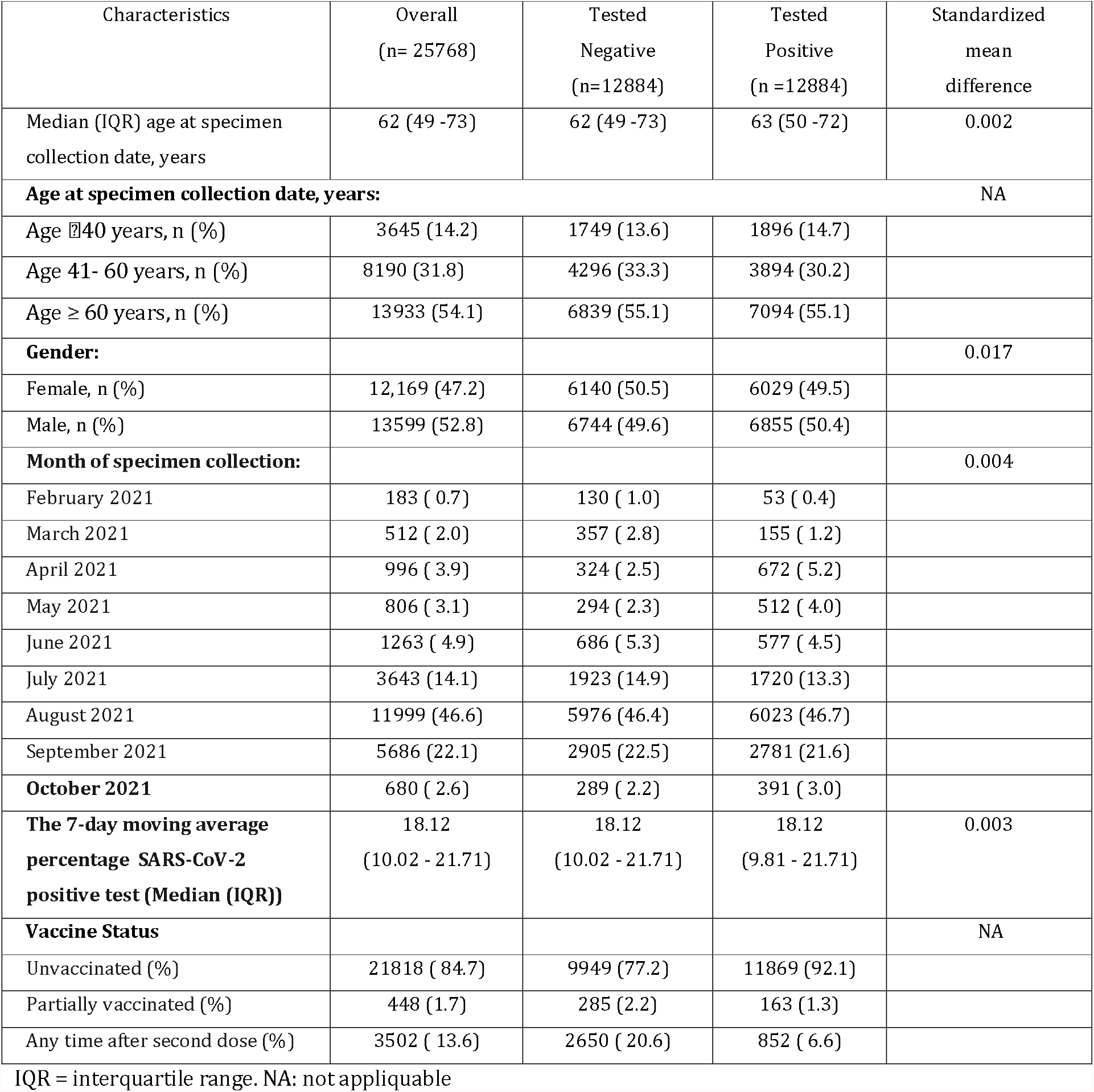
Baseline characteristics of individuals tested positive for SARS-COV-2 infection by rt-PCR test and Propensity Score–Matched control participants who tested negative for SARS-COV-2 infection.

| Characteristics | Overall<br>(n= 25768) | Tested<br>Negative<br>(n=12884) | Tested<br>Positive<br>(n =12884) | Standardized<br>mean<br>difference |
| --- | --- | --- | --- | --- |
| Median (IQR) age at specimen collection date, years | 62 (49 -73) | 62 (49 -73) | 63 (50 -72) | 0.002 |
| **Age at specimen collection date, years:** |  |  |  | NA |
| Age ≤40 years, n (%) | 3645 (14.2) | 1749 (13.6) | 1896 (14.7) |  |
| Age 41- 60 years, n (%) | 8190 (31.8) | 4296 (33.3) | 3894 (30.2) |  |
| Age ≥ 60 years, n (%) | 13933 (54.1) | 6839 (55.1) | 7094 (55.1) |  |
| **Gender:** |  |  |  | 0.017 |
| Female, n (%) | 12,169 (47.2) | 6140 (50.5) | 6029 (49.5) |  |
| Male, n (%) | 13599 (52.8) | 6744 (49.6) | 6855 (50.4) |  |
| **Month of specimen collection:** |  |  |  | 0.004 |
| February 2021 | 183 (0.7) | 130 (1.0) | 53 (0.4) |  |
| March 2021 | 512 (2.0) | 357 (2.8) | 155 (1.2) |  |
| April 2021 | 996 (3.9) | 324 (2.5) | 672 (5.2) |  |
| May 2021 | 806 (3.1) | 294 (2.3) | 512 (4.0) |  |
| June 2021 | 1263 (4.9) | 686 (5.3) | 577 (4.5) |  |
| July 2021 | 3643 (14.1) | 1923 (14.9) | 1720 (13.3) |  |
| August 2021 | 11999 (46.6) | 5976 (46.4) | 6023 (46.7) |  |
| September 2021 | 5686 (22.1) | 2905 (22.5) | 2781 (21.6) |  |
| **October 2021** | 680 (2.6) | 289 (2.2) | 391 (3.0) |  |
| **The 7-day moving average percentage SARS-CoV-2 positive test (Median (IQR))** | 18.12<br>(10.02 - 21.71) | 18.12<br>(10.02 - 21.71) | 18.12<br>(9.81 - 21.71) | 0.003 |
| **Vaccine Status** |  |  |  | NA |
| Unvaccinated (%) | 21818 (84.7) | 9949 (77.2) | 11869 (92.1) |  |
| Partially vaccinated (%) | 448 (1.7) | 285 (2.2) | 163 (1.3) |  |
| Any time after second dose (%) | 3502 (13.6) | 2650 (20.6) | 852 (6.6) |  |

IQR = interquartile range. NA: not applicable

As a function of time after vaccination of second dose vaccination, vaccine effectiveness among persons who had received the second dose 1–30 days earlier was 88% (95% CI, 84-91), 87% (95% CI: 83-90) among those who had received it 31–90 days earlier, 75% (95% CI: 67-80) among those who had received it 91–120 days earlier, 61% (95% CI: 54-67) among those who had received it 121–150 days earlier, 64% (95% CI: 59-69) among those who had received it ≥150 days earlier. The overall vaccine effectiveness any time after the second dose was 73% (95% CI: 71-76). Estimates Vaccine effectiveness against COVID-19 associated severe or critical hospitalisation according to vaccine status and by time since vaccination is shown in Figures 1 and 2.

**Figure 1:**
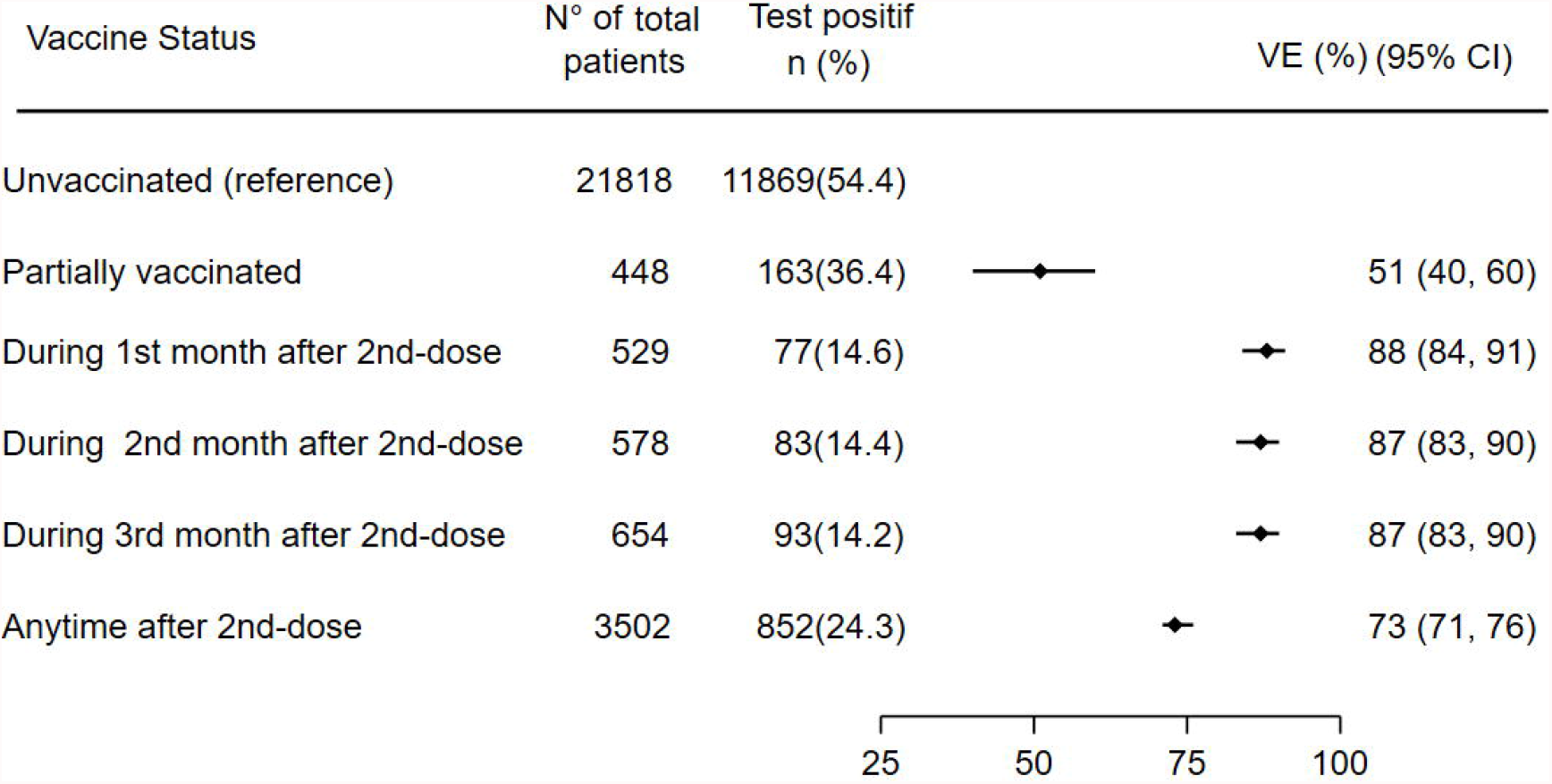
Estimates Vaccine effectiveness of inactivated vaccine BBIBP-CorV against COVID-19-associated severe or critical hospitalisation according to vaccination status and by time since vaccination.

**Figure 2:**
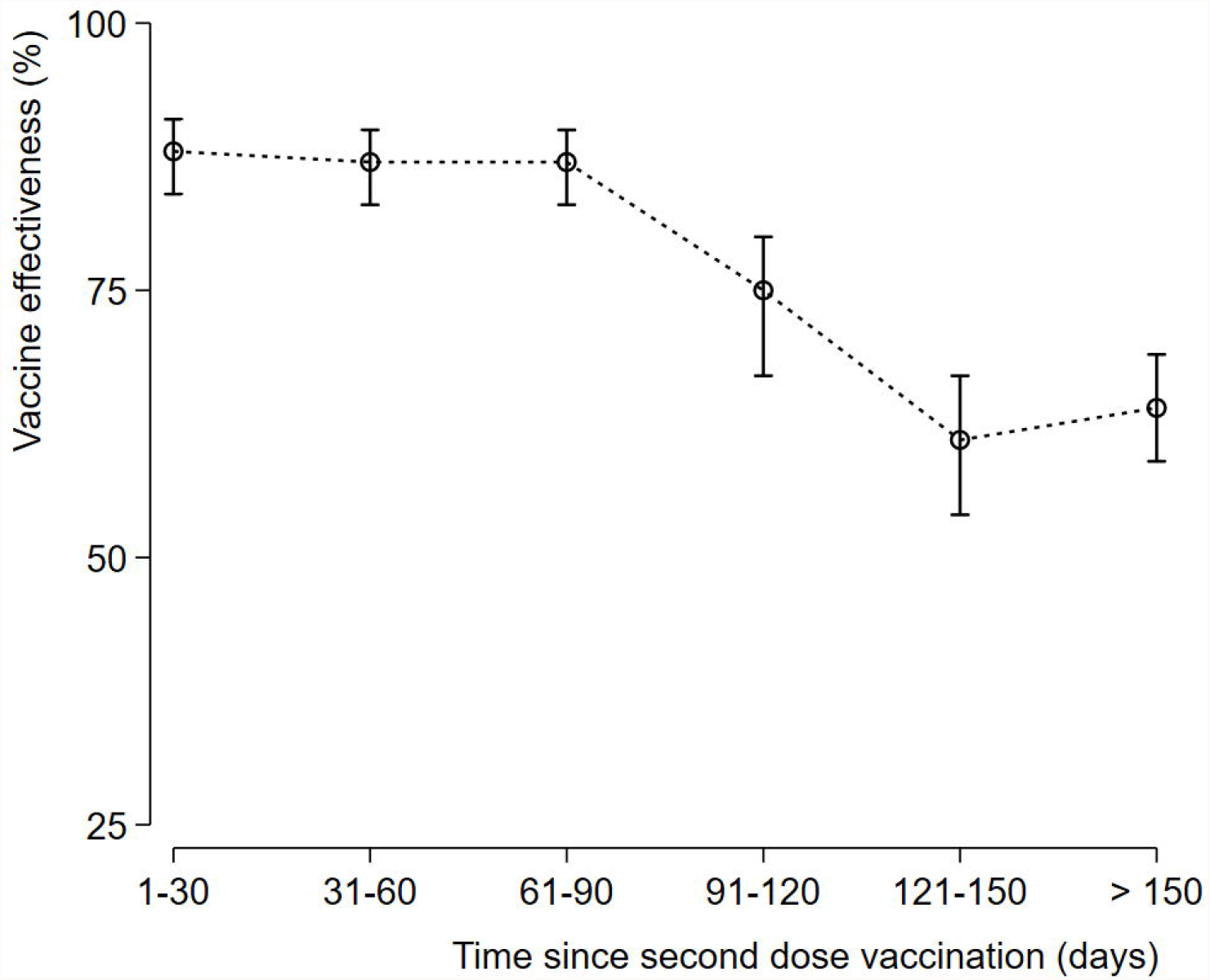
Vaccine Effectiveness of inactivated vaccine BBIBP-CorV against COVID-19- associated severe or critical hospitalisation over time since second dose vaccination. Vaccine Effectiveness at each Time Point: At (1-30) time point ;VE 88% (95% CI: 84-91), At (31-60) time point ; 87% (95% CI: 83-90), At (61-90) time point ; 87% (95% CI: 83- 90), At (91-120) time point ; 75% (95% CI: 67-80), At (121-150) time point ; 61% (95% CI: 54-67), Beyond 150 days time point ; 64% (95% CI: 59-69).

Concerning vaccine effectiveness of inactivated vaccine BBIBP-CorV against COVID-19 associated severe or critical hospitalisation over time since vaccination of the second dose in age subgroup. a) Concerning age subgroup ≥ 60 years old: VE at (1-30) time point was 80% (95% CI: 80-94), at (31-60) time point was 85% (95% CI: 76-91), at (61- 90) time point was 87% (95% CI: 78-92), at (91-120) time point was 64% (95% CI: 48- 75, at (121-150) time point was 53% (95% CI: 42-61), and beyond 150 days’ time point was 66% (95% CI: 59 to 70). b) Concerning age subgroup < 60 yrs old: VE at (1-30) time point was 88% (95% CI: 82 to 92), at (31-60) time point was 90 %(95% CI: 85-93), at (61-90) time point was 90% (95% CI: 85-93), at (91-120) time point was 84% (95% CI: 74-90), at (121-150) time point was 72% (95% CI, 60-80), and beyond 150 days’ time point was 70% (95% CI: 57-79). Any time after the second dose, vaccine effectiveness against severe critical hospitalisation in persons younger than 60 years compared with those aged 60 years and older was: 84% (95% CI: 80-86) and 67% (95% CI: 62-70) respectively. (Figure 3 and 4). In the gender subgroup effectiveness was similar between men and women (figure 4). Estimates of vaccine effectiveness after the second dose as a function of time since second dose vaccination according to subgroup are shown in Figures 3 and 4.

**Figure 3:**
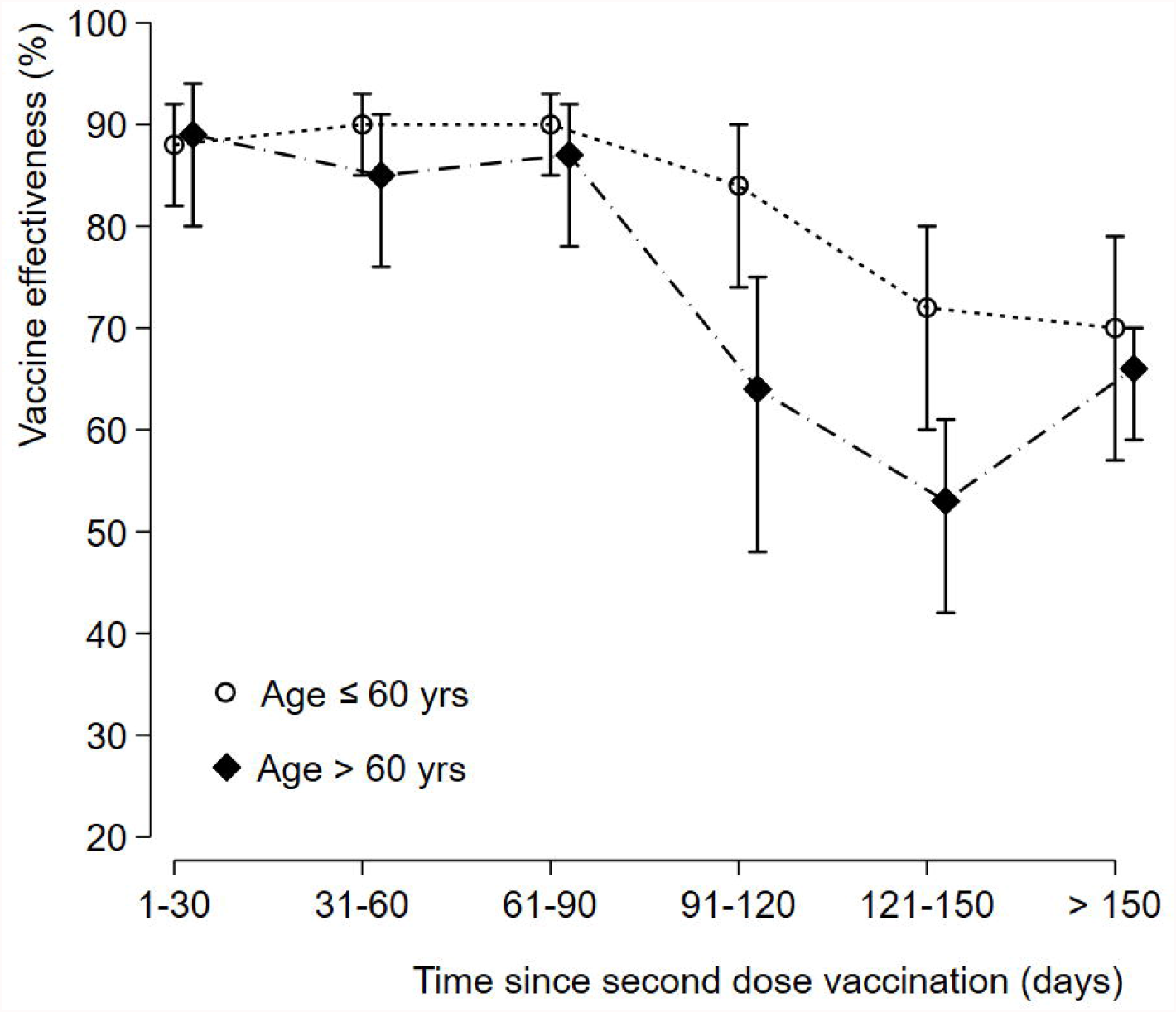
Vaccine effectiveness of inactivated vaccine BBIBP-CorV against COVID-19- associated severe or critical hospitalisation over time since second dose vaccination according to age subgroup. Vaccine Effectiveness at each Time Point in age subgroup ≤ 60 years old: VE at (1-30) time point; 80% (95% CI: 80-94).VE at (31-60) time point; 85% (95% CI: 76-91). VE at (61-90) time point; 87% (95% CI: 78-92). VE at (91-120) time point; 64% (95% CI: 48-75). VE at (121-150) time point; 53% (95% CI: 42-61). VE Beyond 150 days’ time point; 66% (95% CI: 59-70). Vaccine Effectiveness at each time point in age subgroup < 60 yrs old: VE at (1-30) time point; 88% (95% CI: 82-92). VE at (31-60) time point; 90 %(95% CI: 85-93). VE at (61-90) time point; 90% (95% CI: 85- 93). VE at (91-120) time point; 84% (95% CI: 74-90). VE at (121-150) time point; 72% (95% CI: 60-80). VE Beyond 150 days’ time point; 70% (95% CI: 57-79).

**Figure 4:**
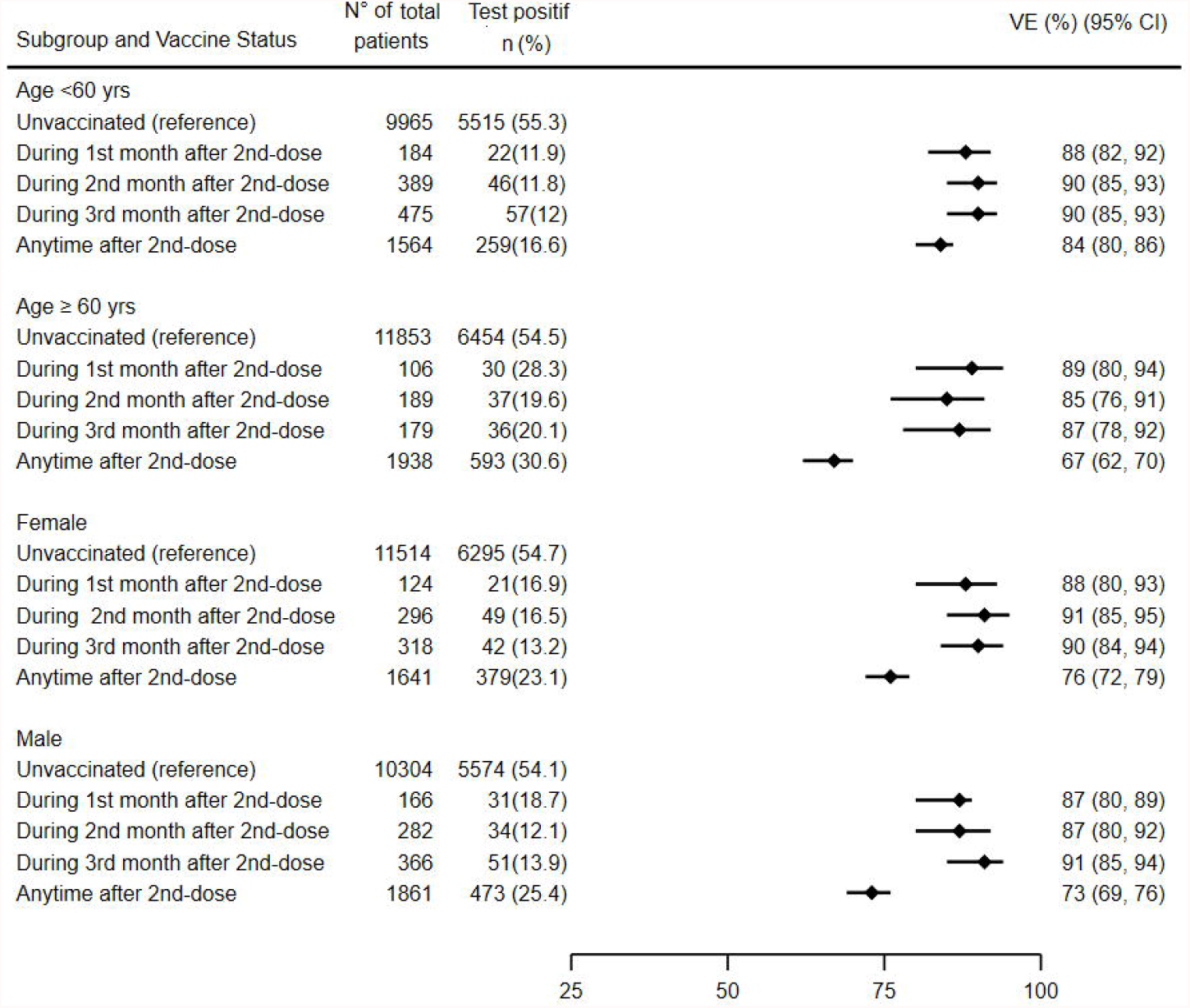
Estimates Vaccine effectiveness of inactivated vaccine BBIBP-CorV against COVID-19-associated severe or critical hospitalisation according to age and gender over time since second dose vaccination.

## DISCUSSION

We provide estimates of the vaccine effectiveness (VE) of the administration of the Sinopharm vaccine in a countrywide mass vaccination campaign for the prevention of severe and critical COVID-19 associated hospitalization in the Kingdom of Morocco. Among fully immunized persons, the vaccine effectiveness at any time during the first 9 months after the second dose was 73%. Due to the longer follow-up time for this study since vaccine rollout, this study provides information on the stability profile of Sinopharm vaccine effectiveness. Estimates of COVID-19 inactivated Sinopharm vaccine effectiveness (VE) decline on the time elapsed since the second dose because of waning vaccine-induced immunity over time, but also by possible increased immune evasion by SARS-CoV-2 variants ^11^or a combination of these and other factors.

Our results showed that the effectiveness of Sinopharm vaccine against severe COVID-19 associated hospitalizations reached a maximum level during the first month, and remains high and durable during the three months after the second dose (87-90%), declines little in the fourth month (75%), and stabilizes around 60% beyond the fifth month.

In a short time since vaccine rollout, our results are consistent with previous studies^12^; but not with the results of phase III clinical trial given the small number of incident severe cases^13^.

The large sample size and long duration of follow-up in this study allowed us to determine the timing of the peak and the duration of protection.

Vaccine effectiveness results differ in age subgroup analysis, with those aged 60 years or older showing an earlier and more evident decline beyond the third-month post-vaccination. Due to immunosenescence and comorbidities, older people are more susceptible to infections and respond less well to vaccination^14^.

Due to the nature of real-world effectiveness studies, they can be subject to selection bias, confounding factors, and missing data, therefore requiring careful study design. A test-negative design case-control study was in the general population, which may be subject to collider bias. Due to the observational study design, selection bias and confounding effects were inevitable limitations. Then, this analytical approach was implemented to reduce potential bias due to variation in the epidemic phase, gradual vaccination^15^ and other confounders^16^. The test-negative design has been used extensively to estimate VE among medically attended influenza virus illness and is believed to minimize biases associated with access to vaccines and healthcare-seeking behaviours^17^, and recommended by the WHO interim guidance^18^.

In conclusion, in accordance with the results of phase III clinical trials, the licensed Sinopharm vaccine is highly protective against SARS-CoV-2 in real conditions. In addition, this study confirms the durability of the protection. VE depends not only on the effectiveness of the vaccine itself but also on factors such as the age of the recipient and time since the latest dose.

This study of vaccine effectiveness focuses on the specific subset of cases - severe or critical - with the greatest burden on health systems, regardless of their level of maturity, and requires the most attention for vaccine effectiveness evaluations. The elderly population is more vulnerable and at greater risk of immune depletion overtime after full immunization and deserves special attention. Therefore, maintaining the stability of their protection, through either continued social distancing or active vaccination, must be a priority. Governments may face declining public confidence in the vaccines used in their country, post-licensure observational evaluations are important to inform policy decisions about vaccine introduction, to optimize vaccine implementation programs and to provide evidence in use support of vaccines and investments by governments and donors, and above all to convey a convincing message to the population. By combining these features, the present study has generated new evidence that helps in making informed decisions.

## Data Availability

All data produced in the present study are available upon reasonable request to the authors

## Author contributions

All authors conceived the study. JB, MO and RR completed analyses with guidance from RA, and JC. JB, MO, RR, and RA curated and validated the data. JB wrote the manuscript. All authors contributed to, and approved, the final manuscript. The corresponding author attests that all listed authors meet authorship criteria and that no others meeting the criteria have been omitted.

## Conflict of interest

None declared.

## Acknowledgment

To the Ministry of Health of the Kingdom of Morocco for providing the Database subject of the study. We also thank the assistance offered by funders and each cooperative institution, and we are very grateful to every medical staff who contributed in collecting data in this epidemic.

## Notes

### Competing Interest Statement

The authors have declared no competing interest.

### Clinical Protocols

https://osf.io/at3yf

### Funding Statement

This study did not receive any funding

### Author Declarations

The Rabat local ethic committee review board for biomedical research at Mohammed V University Decision N/21.

## REFERENCES

1. https://covid19.who.int/region/emro/country/ma (accessedJanuary 18, 2022).

2. https://www.liqahcorona.ma/(accessed January 18, 2022).

3. http://inh.ma/?page_id=431 (accessed January 18, 2022).

4. World Health Organization. COVID-19 clinical management: living guidance https://www.who.int/publications/i/item/WHO-2019-nCoV-clinical-2021-1. (Accessed January 18, 2022).

5. Teerawattananon Y, Anothaisintawee T, Pheerapanyawaranun C, Botwright S, Akksilp K, Sirichumroonwit N, Budtarad N, Isaranuwatchai W. A systematic review of methodological approaches for evaluating real-world effectiveness of COVID-19 vaccines: Advising resource-constrained settings. PLoS One. 2022 11;17 (1): e0261930. doi: 10.1371/journal.pone.0261930.

6. Cohen J. Statistical power analysis in the behavioral sciences. 2nd ed. New York: Routledge, 1988.

7. Imbens, G., & Rubin, D. Estimation du score de propension. En causalité Inférence pour les statistiques, les sciences sociales et biomédicales : une introduction. 2015 (281-308). Cambridge: Cambridge University Press. doi:10.1017/CBO9781139025751.014

8. Estimating COVID-19 vaccine effectiveness against severe acute respiratory infections (SARI) hospitalisations associated with laboratory-confirmed SARS-CoV-2: an evaluation using the test-negative design: guidance document. WHO-EURO-2021-2481-42237-58308-eng.pdf (1.185Mo) (accessed January 18, 2022).

9. Austin PC. Type I error rates, coverage of confidence intervals, and variance estimation in propensity-score matched analyses. Int J Biostat. 2009;5 doi:10.2202/1557-4679.1146

10. Hightower AW, Orenstein WA, Martin SM. Recommendations for the use of Taylor series confidence intervals for estimates of vaccine efficacy. Bull World Health Organ. 1988; 66:99–105.

11. Wall EC, Wu M, Harvey R, et al. Neutralising antibody activity against SARS-CoV-2 VOCs B.1.617.2 and B.1.351 by BNT162b2 vaccination. Lancet 2021;397:2331–

12. https://doi.org/10.1016/S0140-6736(21)01290-3

13. Ara A, Undurraga EA, González C, Paredes F, Fontecilla T, Jara G, et al. Effectiveness of an Inactivated SARS-CoV-2 Vaccine in Chile. N Engl J Med. 2021; 2; 385 (10):875–884. doi: 10.1056/NEJMoa2107715.

14. Al Kaabi N, Zhang Y, Xia S, et al. Effect of 2 Inactivated SARS-CoV-2 Vaccines on Symptomatic COVID-19 Infection in Adults: A Randomized Clinical Trial. JAMA. 2021 ; 326(1):35–45. doi:10.1001/jama.2021.8565

15. Brosh-Nissimov T, Orenbuch-Harroch E, Chowers M, Elbaz M, Nesher L, Stein M, et al. BNT162b2 vaccine breakthrough: clinical characteristics of 152 fully vaccinated hospitalized COVID-19 patients in Israel. Clin Microbiol Infect. 2021; 27(11):1652–1657. doi: 10.1016/j.cmi.2021.06.036.

16. Jacoby, P. & Kelly, H. Is it necessary to adjust for calendar time in a test-negative design? Responding to: Jackson ML, Nelson JC. The test-negative design for estimating influenza vaccine effectiveness. Vaccine 2013;31(17):2165–8

17. Pearce, N. Analysis of matched case–control studies. BMJ (2016).352, i969

18. Jennifer R. Verani, Abdullah H. Baqui, Claire V. Broome, Thomas Cherian, Cheryl Cohen, Jennifer L. Farrar, et al Case-control vaccine effectiveness studies: Preparation, design, and enrollment of cases and controls, Vaccine,2017, 3295–3302, https://doi.org/10.1016/j.vaccine.2017.04.037.

19. Évaluation de l’efficacité du vaccin COVID-19 (Directives provisoires) Genève, https://www.who.int/publications/i/item/WHO-2019-nCoV-vaccine_effectiveness-measurement-2021.1 (Accessed January 18, 2022).

